# Fronto-parietal network dynamics to understand deficits in attention performance in multiple sclerosis

**DOI:** 10.1101/2023.07.08.23292404

**Authors:** Thomas Welton, Dewen Meng, Roshan das Nair, Cris S Constantinescu, Dorothee P Auer, Rob A Dineen

**Affiliations:** Radiological Sciences, Division of Clinical Neuroscience, University of Nottingham, W/B 1441, Radiological Sciences, Queen’s Medical Centre, Derby Road, Nottingham, NG7 2UH, UK; Sir Peter Mansfield Imaging Centre, University of Nottingham, W/B 1441, Radiological Sciences, Queen’s Medical Centre, Derby Road, Nottingham NG7 2UH, UK; National Neuroscience Institute, 11 Jln Tan Tock Seng, Singapore 308433; Institute of Mental Health, Jubilee Campus, University of Nottingham Innovation Park, Triumph Road, Nottingham NG7 2TU, UK; Division of Psychiatry and Applied Psychology, University of Nottingham, Jubilee Campus, University of Nottingham Innovation Park, Triumph Road, Nottingham NG7 2TU, UK; Clinical Neurology, Division of Clinical Neuroscience, University of Nottingham, C Floor, South Block, Queen’s Medical Centre, Nottingham, NG7 2UH, UK; NIHR Nottingham Biomedical Research Centre, C Floor, South Block, Queen’s Medical Centre, Nottingham, NG7 2UH, UK; Duke-NUS Graduate Medical School, Singapore

**Keywords:** functional connectivity, multiple sclerosis, magnetic resonance imaging, diffusion imaging, attention, cognition, networks

## Abstract

**Introduction:** Impaired attention performance is a significant burden to people with multiple sclerosis (MS). Brain connectivity fluctuates with transitions between cognitive states, so measurement of network dynamics during these conditions may help to understand MS-related attention impairment.

**Methods:** In people with MS and healthy controls, attention was measured using the Attention Network Test. 3T MRI was used to measure structural connectivity and both static and dynamic functional connectivity in the attention-related fronto-parietal network (FPN) at rest and during an attentionally-demanding task. Groups were compared on connectivity of the FPN during rest and task performance. Relationships between network connectivity and attention performance were tested using linear regression.

**Results:** The sample comprised 37 people with MS and 23 matched controls. At rest, people with MS had significantly lower structural connectivity (R^2^=0.13, p=0.004), lower static functional connectivity (R^2^=0.07, p=0.032) and higher dynamic functional connectivity (R^2^=0.08, p=0.026) of the FPN. Higher dynamic connectivity was significantly associated with poorer attention performance in people with MS (R^2^=0.20, p=0.008). During attention-task performance, static functional connectivity was greater in people with MS than controls (R^2^=0.10, p=0.008). The task-induced reduction in static connectivity (relative to rest) was directly related to attention performance (R^2^=0.23, p<0.001).

**Conclusion:** Increased dynamic functional connectivity of the FPN at rest may be a useful indicator of deficits in sustained attention in people with MS. The transition from rest to active-attentive state is accompanied by an increase in dynamic connectivity, and decrease in static connectivity which may be helpful in understanding aetiology and treatment of attention impairment.

## Introduction

Cognitive impairment occurs in 40-65% of people with multiple sclerosis (MS) and frequently involves a reduced capacity for sustained attention or concentration [1–3]. Attention impairments are reported at rates between 20% and 50%, depending on the task used [4] and greatly impact patients’ quality of life [5]. Specifically, sustained attention abnormalities are often associated with difficulties in behavioural, learning, emotional, and cognitive aspects especially during adolescence [6] and sustained attention tasks are useful for quantifying cognitive fatigue [7]. Evidence for the effectiveness of treatments for cognitive impairment is equivocal [8] which, may, in-part, be due to difficulty in measuring cognitive symptoms. Neuroimaging may offer a non-invasive, direct and quantitative way to measure cognition-related neural changes in MS.

Neuroimaging studies have shown relationships between imaging measures and attention impairment in people with MS; for example, grey matter atrophy [9], abnormal white matter diffusivity [10] and greater lesion volumes [11]. However, normal brain structure and function is now understood to depend on the interconnectivity of anatomical regions or networks [12]. Accordingly, attention performance may be based on the connectivity of a set of attention-related areas [13–16], which includes the superior and middle frontal gyri, posterior cingulate, superior parietal lobule, intraparietal sulci, superior temporal gyri and lateral occipital gyri, known as the fronto-parietal network (FPN). Studies show that functional connectivity of these networks is disrupted in MS [17]; and in other cognitive networks such as a memory-related network to the n-back task [18], in motor networks [19] and visual networks [20]. However, the specific nature of the FPN disruption and how it relates to attention performance is not fully understood.

Dynamic functional connectivity, i.e. the variation in strength of connectivity between brain regions over time, is a recently-described technique for analysis of fMRI data, and may be important for understanding cognition in MS [21]. Dynamic functional connectivity analyses include “sliding window” (the simplest approach, measuring the variance in connectivity at the scale of seconds to minutes), “time-frequency” (incorporating information from the frequency domain at shorter timescales, for example using the wavelet transform), “point process” (measuring single time-point activations) and “graph theoretic” (variation in holistic topological measures of network integration and segregation) analyses [22]. There are now many examples of the application of these techniques in neurological disease [23]; for example abnormal default-mod sub-network “dwell times” in Alzheimer’s [24] and Parkinson’s disease [25]. Studies of resting dynamic connectivity in MS have also shown differences in state-dwelling times in cognitively impaired patients [26], reduced hippocampal dynamic connectivity related to maintained memory performance [27], greater dynamic connectivity of specific connection pairs related to lower lesion loads [28] and reduced network dynamics in cognitively impaired patients [29], mostly using the simple sliding window dynamic connectivity approach. There are also EEG studies showing patient-control differences in network dynamics during attentional tasks [30–32]. The dynamic functional connectivity approach may be useful in investigating attention deficits because attentional focus inherently varies in its target and strength based on demands [33, 34] and therefore may require a tool that can detect these variations [35].

Across this body of research, major knowledge gaps exist. Namely, in understanding how dynamic functional connectivity differs between intrinsic and stimulated states. In MS, no study has contrasted brain dynamics during rest with that during task performance which, according to research in other patient populations showing functionally relevant variations in static and dynamic connectivity between resting and task states, may be important in understanding the link to cognition [36, 37]. It is implied that network specific activation during task performance leads to an increase in functional connectivity and decrease in dynamic connectivity as the network becomes driven by the task rather than interference across other activities. However, this link is complex and likely non-linear because a deficit phase may show different associations than a compensatory phase. One possibility is that network-specific activation during task performance leads to an increase in functional connectivity and decrease in dynamic connectivity as the network signal becomes driven more by the task than interference across other activities.

We propose that dynamic functional connectivity of the FPN may enable a better understanding of mechanisms behind attention performance and its impairment in people with MS by showing the direction and magnitude of changes in an attention-related network (FPN) between groups and between states. We will apply the sliding-window variance technique to quantify dynamic functional connectivity in an attention-related network, as the most standardised, widely-used and reproducible method from the literature. We hypothesised that functional connectivity of an attention-related network during task performance would (a) be abnormal in people with MS compared to controls, with people with MS having lower static but higher dynamic connectivity, (b) correlate with measures of attention performance and alter under sustained attentionally-demanding task performance compared to rest, with better task performance relating to greater static connectivity and reduced dynamic connectivity, and (c) be different between anatomical regions with a known role in attention and whole-brain connectivity measures.

## Methods

### Subjects

Participants were recruited from MS clinics at a large UK National Health Service hospital according to the following criteria: aged 18-65 years, clinically-definite relapsing-remitting MS (RRMS) or secondary-progressive MS (SPMS), no other neurological or psychiatric conditions and no relapses or changes in medication 30 days prior (sample size calculation in Supplemental Information). RRMS and SPMS were diagnosed according to the McDonald criteria [38]. We also recruited age-, gender- and education-matched healthy participants via recruitment posters, who also were aged 18-65 and had no neurological or psychiatric disease. All participants underwent a 2-hour visit for cognitive testing and MRI between September 2014 and January 2017 and gave written informed consent. The study was approved by the Nottingham Research Ethics Committee 2 (14/EM/0064).

### Neurocognitive Testing

Participants underwent testing for attention performance using the Attention Network Test (ANT) [39]. The ANT is a computer based test which shows a sequence of visual stimuli which can be used to predict the proceeding action. ANT measures three subscales based on reaction times resulting from a warning signal (“alerting”), in response to a cue about where the target will appear (“orienting”) and with correct directional response of a central marker (“executive”) [40]. The ANT was used as the primary measure for attention in all statistical tests, while two other secondary measures were also tested.

Secondary cognitive measures that include a significant attention component were the Paced Auditory Serial Addition Test (PASAT-3) [41] and Symbol Digit Modalities Test (SDMT) [42]. We also measured known confounding factors: fatigue using the Modified Fatigue Impact Scale (MFIS) [43], depression using the Beck Depression Inventory (BDI-II) [44] and sleep quality using the Pittsburgh Sleep Quality Index (PSQI) [45]. For further characterisation of the sample and ease of comparison to other studies, we also recorded the MS Functional Composite (MSFC) [46] which includes Nine-Hole Peg Test (9HPT) and Timed 25-Foot Walk (T25FW). The cohort was naïve to all tests except those in the MSFC.

### Magnetic Resonance Imaging

MRI was performed on a 3T GE Discovery 750 scanner (General Electric Healthcare; Milwaukee, WI) with 32-channel head coil. The protocol included T1-weighted fast-spoiled gradient echo (FSPGR; FA=8°, matrix 256×256×156, voxel size 1×1×1mm, TE=3.17ms, TI=900ms, TR=8200ms, FOV=256mm, NEX=1), T2-weighted fluid attenuated inversion recovery (FLAIR; FA=111°, TE=120ms, TR=8000ms, TI=2250ms, matrix 512×512×46, voxel size 0.46×0.46×3mm, FOV=235.5mm), diffusion MRI (dMRI; four b=0 volumes and 32 diffusion weighted volumes at b=1000, matrix 128×128×66, voxel size 2mm^3^) and functional MRI (fMRI; FA=80°, TE=36ms, TR=2200ms, matrix 64×64×37, voxel size 3.75mm^3^) at rest (9 minutes, 245 volumes) and during task performance (6 minutes, 165 volumes).

### fMRI Task Paradigm

Subjects performed a 6-minute sustained attention task while in the scanner. The task, the modified SDMT (mSDMT), was designed to induce cognitive load by continuous sustained attention and information processing [47], and was modelled on the SDMT but adapted for use in the MRI scanner to allow responses using buttons (Figure 1a). We implemented the task in E-Prime (version 2.0; Psychology Software Tools; Pittsburgh, PA). Error rates were recorded and used to quantify performance.

**Figure 1.**
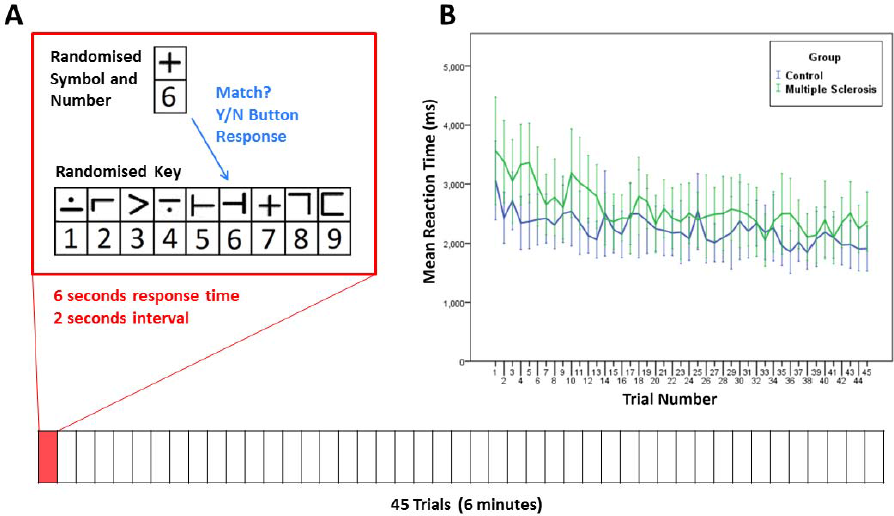
Design of fMRI task and reaction times. The task comprised 45 trials, each 6 seconds with 2-second intervals. During each trial, the subject decides whether the bottom symbol pair matches any of the presented randomised pairs and responds by pressing a button (“match” or “no-match”).

### Parcellation and Fronto-Parietal Network Definition

T1-weighted images were segmented using FreeSurfer (version 5.3; http://surfer.nmr.harvard.edu/) to generate sets of 164 grey matter ROIs based on the Destrieux atlas [48]. We defined the FPN based on literature [13, 49] and the NeuroSynth meta-analytic topic map for attention control (neurosynth.org; topic 264, version 5, July 2018, [50]). We selected the relevant labels from the FreeSurfer cortical parcellation, including the superior and middle frontal gyri, posterior cingulate, superior parietal lobule, intraparietal sulcus, superior temporal gyrus and lateral occipital gyrus bilaterally (Figure 2). We also gathered whole-brain volumes from FreeSurfer.

**Figure 2.**
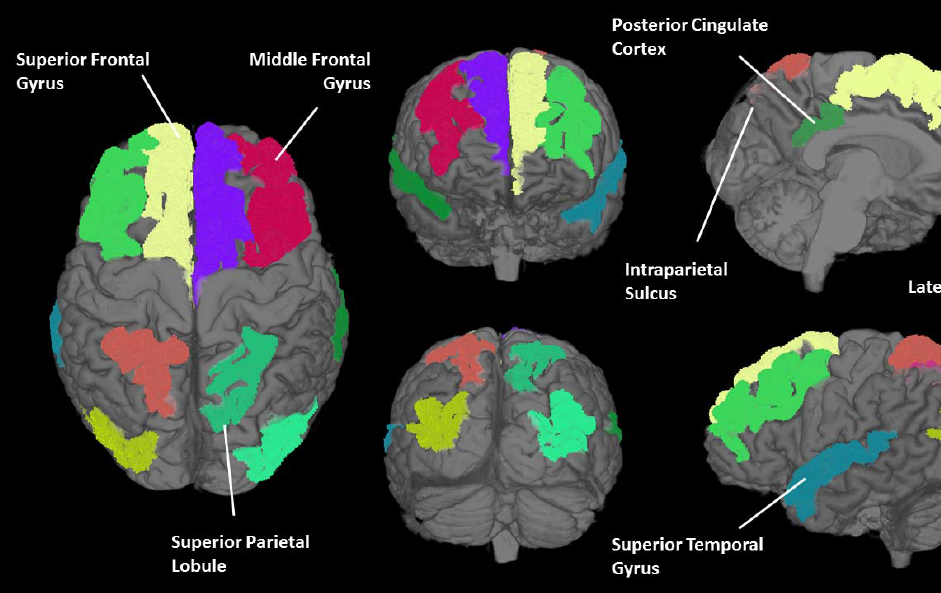
Regions of the fronto-parietal network in an example subject.

### Lesion Volumes

The volume of T2-hyperintense lesions was measured on the T2-FLAIR images using JIM (version 5.0; Xinapse Systems; Essex, UK) for fuzzy semi-automatic lesion segmentation. A single point was manually placed in each lesion and the JIM software then automatically expands a region until the lesion is filled in a 3D mask for further use in segmentation/registration. Lesion volumes were normalised to whole-brain volume.

### Static Functional Connectivity

Quality of the fMRI data was ascertained using data quality best practices from the fBIRN initiative [51] including inspection of power spectra, implemented in MATLAB (version 2013a; TheMathWorks; Natick, MA). fMRI data were pre-processed in FSL MELODIC [52] using the following steps: (1) removal of the first 5 volumes, (2) motion correction and (3) spatial smoothing with a 5mm Gaussian kernel. Regions with a timeseries average greater than 3 standard deviations below the mean were ignored in the analysis. Outlying timepoints were detected using the FSL motion outliers tool [52] and missing data points replaced using cubic interpolation. High-pass filtering with a 1-second cut-off was applied. Spatial ICA was performed using MELODIC [53] to manually identify (following the steps provided by Griffanti, Douaud [54]) and filter-out (using the “fsl_regfilt” tool) components consisting predominantly of noise, with no limit on the number of components. To apply the anatomical regions to the fMRI data, we first registered the task fMRI images to the resting-state fMRI images. We then created a transform from the resting-state fMRI to the T1-weighted image using the boundary-based registration tool in FLIRT [55]. This transformation was applied in reverse (and, for the task fMRI, the task-to-rest transformation) to the anatomical regions to produce personalised grey-matter atlases in fMRI space. The transformed masks were eroded by one voxel to reduce partial volume effects and contributions from white matter or CSF. The mean timeseries from each region was imported to MATLAB (R2017b; Natick, Massachusetts, USA) and static functional connectivity quantified as the Pearson correlation coefficient between each pair of regions in the FPN (7-by-7 matrix). As a control, the same was performed for all regions in the FreeSurfer parcellation (164-by-164). This measure was chosen to reflect the level of activity within the network.

### Dynamic Functional Connectivity

Using the same timeseries data as above, dynamic functional connectivity of the FPN was measured during rest and task performance using the sliding window approach of previous studies [27, 56-58]. In short, the fMRI region timeseries were split into overlapping windows of 59.4-second length (27 TRs) and an 8.8-second shift (4 TRs), resulting in 55 windows for the resting-state fMRI and 35 for the task fMRI. Within each window, static functional connectivity was calculated. The absolute difference between consecutive windows was calculated and summed across all windows. With this approach, the resulting 7-by-7 matrix reflected the amount of variance in functional connectivity over time.

### Structural Connectivity

Diffusion data were processed using FSL Diffusion Toolbox and probtrackx2 [59]: first, eddy-current induced distortions and subject motion were corrected using Eddy [60]. All image volumes were registered to the average b=0 image and non-brain structures were removed. Diffusion parameters were estimated and ROIs from the two atlases were registered to diffusion space by registering the T1 brain image to the average b=0 image by rigid transformation and applying this transformation to the atlas. Fibre tracking was applied to every pair of ROIs to create matrices of the number of reconstructed streamlines between them, from the seeded 5,000. This was measured twice (A-to-B and B-to-A). The following standard formula was then applied in order to sum the streamline counts for each direction and scale the result by their volumes [61]: *a_ij_* = (*s_ij_* + *s_ji_*)⁄(*m_i_* + *m_j_*). Where a_ij_ is the corrected summary streamline count, s_ij_ and s_ji_ are streamline counts in each direction between ROIs i and j, and m_i_ and m_j_ are voxel counts in each ROI. Diffusion data were incomplete for 4 participants from the MS group.

### Statistical Analysis

All variables were first tested for normality using Kolmogorov-Smirnov tests and by visual inspection of histograms. The level of statistical significance for all tests was set at 5%. Group differences in age, sex, education and head motion were assessed by chi-square test or t-test.

To test for differences in FPN volumes between groups and between hemispheres, and for differences in lesion and whole-brain volumes, we applied two-sample t-tests (Supplemental Information).

To test for between-group differences in FPN connectivity, we used multiple regression, inputting the dummy group variable as the dependent variable, age, gender, education and average head motion as a block of independents via the enter method, and the connectivity measure in a second block. For tests in the FPN, using either dynamic or static connectivity, and structural connectivity, this comprised 5 tests. We evaluated the impact of multiple comparisons against a Bonferroni-corrected alpha (α′=0.05/5=0.01).

Relationships between brain connectivity measures in the FPN and attention performance (ANT) were assessed by linear regression in the MS group. For these tests, FPN average measures of static functional connectivity, dynamic functional connectivity and structural connectivity were normalised to their whole-brain equivalents, so that they did not simply reflect global inter-subject differences. The primary analysis was of the relationship between the ANT Alerting score [62, 63] and dynamic connectivity in the FPN at rest. We also tested the relationship to static functional and structural connectivity. Again, we used a Bonferroni-corrected alpha for 3 tests to account for multiple comparisons (α′=0.05/3=0.017). Last, we calculated the task-induced change in static and dynamic functional connectivity as the difference between resting and task-active connectivities and tested its relationship to attention performance (ANT) using linear regression. For all tests, age, gender, education, average head motion and normalised lesion volume were entered as covariates (via the backward-stepwise method).

## Results

### Demographics and Neurocognitive Performance

Thirty-seven people with MS and 23 healthy controls were recruited (Table 1). The groups were not significantly different in age, sex, education or head motion. The MS group performed significantly worse in all tests of attention (ANT, SDMT, PASAT), depression, fatigue and sleep quality. Compared to normative MS data, our sample performed poorly in attention but was similar in terms of physical disability, sleep quality or depression (Table S1). MS patients had significantly greater lesion volumes and smaller brain volumes than controls (Supplemental Information).

**Table 1.** Demographic and cognitive data. Data reported as mean ± standard deviation. Group difference was tested using two-tailed independent-samples t-test except where indicated.

|  |  |  | **Group Difference** |  |
| --- | --- | --- | --- | --- |
|  | **Multiple Sclerosis** | **Control** | **Test statistic (t)** | **p** |
| **N** | 37 | 23 |  |  |
| **Age (years)** | 48±11 | 42±12 | 1.92 | 0.060 |
| **Sex (%F)** | 81 | 73 | 0.43 <sup>a</sup> | 0.735 |
| **Years of Full-Time Education after age 16 (years) <sup>b</sup>** | 2±2 | 3±2 | -1.69 | 0.100 |
| **Handedness (%R)** | 79 | 87 | 0.112 <sup>a</sup> | 0.743 |
| **MS Subtype** | RR=22, SP=15 |  |  |  |
| **Age of Onset (years)** | 31±6 | | | |
| **Disease Duration (years)** | 17±10 |  |  |  |
| **Head Motion During fMRI (mean relative displacement, mm)** | 0.159±0.102 | 0.125±0.051 | 1.48 | 0.146 |
| **TWMLL (mL)** | 7.77±8.87 | 0.42±0.87 | 3.95 | <0.001 ** |
| **MSFC** | -0.38±0.69 | 0.64±0.43 | -6.30 | <0.001 ** |
| **PASAT** | 36.31±2.44 | 44.87±2.43 | -2.22 | 0.030 * |
| **9HPT** | 25.03±7.09 | 22.23±6.71 | -6.18 | <0.001 ** |
| **T25FW** | 7.5±3.3 | 4.6±0.8 | 1.47 | 0.150 |
| **PSQI** | 8.35±3.42 | 4.50±1.41 | 3.05 | 0.003 ** |
| **MFIS** | 54.25±19.98 | 11.00±10.08 | 9.43 | < 0.001 ** |
| **BDI** | 18.21±12.37 | 2.00±3.22 | 4.22 | < 0.001 ** |
| **SDMT** | 41.91±13.09 | 57.00±10.74 | -4.56 | < 0.001 ** |
| **ANT Alerting** | -0.97±14.53 | 17.33±18.57 | -4.05 | < 0.001 ** |
| **ANT Orienting** | 20.57±24.19 | 36.29±23.95 | -2.34 | 0.023 * |
| **ANT Conflicting** | 99.12±49.05 | 170.91±72.45 | -4.35 | < 0.001 ** |

MS = multiple sclerosis, fMRI = functional magnetic resonance imaging, TWMLL = total white matter lesion load, MSFC = multiple sclerosis functional composite, PASAT = paced auditory serial addition task, 9HPT = nine hole peg test, T25FW = timed 25-foot walk, PSQI = Pittsburgh sleep quality index, MFIS = modified fatigue impact scale, BDI = beck depression inventory, SDMT = symbol digit modalities test, ANT = attention network test.
<sup>a</sup> Chi-square
<sup>b</sup> Post-compulsory education in the UK (16 years of age).
\* p<0.05
\*\* p<0.01

### Group Differences within Fronto-Parietal Network Connectivity at Rest

While at rest, there were moderate strength significant differences between the MS and control groups in terms of static and dynamic functional connectivity within the FPN (Table 2; static: F=2.91, R^2^=-0.07, p=0.032; dynamic: F=3.28, R^2^=0.08, p=0.026), with MS patients having lower static connectivity (Figure 3a) and higher dynamic connectivity. Structural connectivity (Figure 3b) was also significantly lower in the FPN in people with MS (Figure 3b; F=8.79, R^2^=-0.13, p=0.004) and this result persisted after Bonferroni correction, as was grey matter volume (Figure 3c). Across the whole brain, there was no significant effect for people with MS having lower structural connectivity (F=2.24, R^2^=0.07, p=0.063) or for functional connectivity (either static or dynamic; F=*0.34*, R^2^=0.01, p=0.551; F=0.47, R^2^=0.02, p=0.495).

**Table 2.** Comparisons between MS and control groups on brain connectivity measures at rest.

| *Imaging Measure* | *F* | *p* | *R<sup>2</sup>* |
| --- | --- | --- | --- |
| ***Fronto-parietal network*** |  |  |  |
| *Resting Static FC* | *2.911* | *0.032 \** | *0.074* |
| *Resting Dynamic FC* | *3.276* | *0.026 \** | *0.079* |
| *Structural Connectivity* | *8.791* | *0.004 \*\** | *0.134* |
| ***Whole Brain*** |  |  |  |
| *Resting Static FC* | *0.340* | *0.551* | *0.013* |
| *Resting Dynamic FC* | *0.473* | *0.495* | *0.015* |
| *Structural Connectivity* | *2.241* | *0.063* | *0.068* |

MS = multiple sclerosis, FC = functional connectivity.
\* $p<0.05$
\*\* $p < 0.01$

### Relationships between Connectivity of the Fronto-Parietal Network and Attention Performance in MS

Within the MS group, higher dynamic functional connectivity measured in the FPN during rest was significantly related to poorer performance in the ANT Alerting score (Figure 3d; F=3.87, p=0.008, R^2^=-0.20). In this model, age, gender and education were not individually predictive but greater lesion volumes were. Neither corresponding test for static functional connectivity nor structural connectivity was significant (Table 3). The corresponding tests in the whole brain were also non-significant. Of the secondary attention measures, both the PASAT and SDMT scores were related to dynamic FPN functional connectivity in the same direction as for the ANT, but these effects were non-significant when controlling for multiple comparisons and neither were significantly related to static functional connectivity (Table S4).

**Figure 3.**
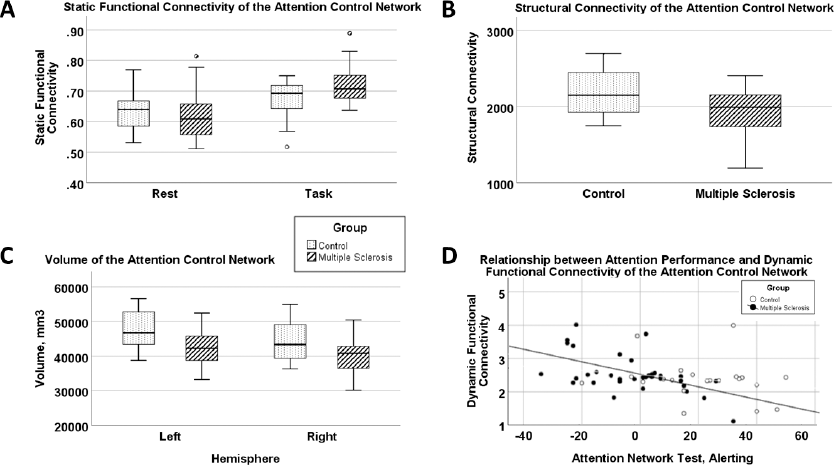
Results of statistical tests on attention related network. (A) comparison of static functional connectivity between resting and task-active states in the fronto-parietal network, (B) comparison between groups of structural connectivity strength in the fronto-parietal network (C) hemispheric volumes of the fronto-parietal network, and (D) dynamic connectivity of the fronto-parietal network compared to attention performance in MS (black dots, R^2^=0.2) and control (white circles).

**Table 3.** Relationships between attention performance and connectivity of the fronto-parietal network at rest in MS.

| *Imaging Measure* | *F* | *p* | *R<sup>2</sup>* |
| --- | --- | --- | --- |
| ***Fronto-parietal network*** |  |  |  |
| Resting Static FC | 2.731 | 0.055 | 0.122 |
| Resting Dynamic FC | 3.870 | 0.008 * | 0.200 |
| Structural Connectivity | 0.424 | 0.698 | 0.039 |
| ***Whole Brain*** |  |  |  |
| Resting Static FC | 1.120 | 0.356 | 0.004 |
| Resting Dynamic FC | 0.384 | 0.735 | 0.000 |
| Structural Connectivity | 1.371 | 0.233 | 0.063 |

MS = multiple sclerosis, FC = functional connectivity.
\* $p < 0.05$
\*\* $p < 0.01$

### Fronto-Parietal Network Connectivity During a Performance of a Sustained Attention Task

Reaction times decreased over the course of the task for both groups (Figure 1b) and reaction times of the MS group were consistently slower than those of controls (two-sample t-test of mean reaction times; control mean: 2240ms, SD 837; MS mean: 2572ms, SD 999; t=-6.522, p=<0.001). There was no difference in the rate of change in RT between groups (two-sample t-test of the slope of the linear regression line; control mean: −13.77, SD 7.27; MS mean: - 18.91, SD 13.45; t=1.326, p=0.196). The proportion of incorrect answers was low for both groups, with the MS group giving 59 incorrect responses out of 1485 trials (4%) and the control group giving 21 incorrect responses out of 1035 trials (2%).

During performance of the continuous sustained attention task, static functional connectivity of the FPN was significantly greater in the MS group than in controls (F=7.65, R^2^=0.10, p=0.008; Table S5; Figure 3a). When comparing connectivity measures during rest against those during task performance in the MS and control groups separately, we found that dynamic functional connectivity of the FPN was significantly lower during task performance than at rest in both groups (Table S2). This “task-induced decrease” in static connectivity (i.e., the difference between rest and task-active states) was significantly related to attention performance in the ANT Alerting score (both groups together; Table S3; F=9.12, p<0.001, R^2^=0.23), with better attention performance relating to a greater rest-to-task increase in static connectivity.

## Discussion

Using functional MRI data at rest and during task performance, and a detailed attention assessment, we found that the FPN was more weakly (lower static connectivity) and more variably interconnected (higher dynamic connectivity) in people with MS, and that this corresponded to poorer attention performance at the group level. During performance of an attentionally-demanding task, the MS group had, conversely, greater synchronicity (static connectivity) of brain regions belonging to the FPN (frontal gyri, posterior cingulate, superior parietal lobule, intraparietal sulci, superior temporal gyri and lateral occipital gyri) compared to controls yet performed worse in the task overall. We then showed that task-induced changes in static functional connectivity explained deficits in attention performance better than either static or dynamic functional connectivity during rest or task alone. These findings appeared to differ between the FPN and whole brain. Together, these findings support the hypothesis that MS-related functional deficits in attention are associated with greater resting dynamic functional connectivity of the attention related FPN.

Static functional connectivity represents the amount of synchronicity between regions of the brain and is thought to reflect connection strength [64]. In our study, static functional connectivity of the FPN was significantly lower at rest in MS compared to controls but higher during task performance, and its change between resting and stimulated states was related to attention performance. This change between resting and task states may reflect functional topological network reorganisation [65]; and may be abnormal in MS due to greater metabolic requirements during the task needed to achieve the same level of maintained performance as controls [66] (as shown by the same rate of change in reaction times).

In contrast, dynamic functional connectivity represents the variability of synchronised cortical metabolic demand and, therefore, has been described as underlying a capability to “switch” between intrinsic and stimulated states [67]. In our sample, higher dynamic connectivity of the FPN at rest was related to poorer attention performance in people with MS, suggesting that a state-changing capability of these regions is functionally important. However, it did not appear to be abnormal compared to our control sample during task performance, nor was its change when switching from rest to task states, whereas static connectivity was. This may suggest that variability in dynamic functional connectivity may be less important for attention performance than static functional connectivity.

A potential limitation of this work is in the heterogeneity of our sample, which included both relapsing-remitting and secondary-progressive patients, and a range of medications. We did not distinguish people with MS based on current medications due to limited statistical power. Age could feasibly be an important confound in this type of study due to differing baseline tissue characteristics [68]. We also were not able to include as many controls in the study as we had estimated would be required, having 4 fewer participants than shown in our power calculation. While the mSDMT task performed inside the scanner was designed to replicate the cognitive process used during the normal SDMT, these two tasks differ in their formats and thus cannot be directly compared, nor can the mSDMT with the ANT. The handedness of the subjects was not homogenous, and so the fMRI findings could have been impacted by the inclusion of both left- and right-handed subjects together. The tractography method used did not take into account the effect of lesions and may be unreliable, and so could be improved by the use of other tools, e.g. MRTrix3 ACT [69, 70]. Finally, we acknowledge that the statistical power of the study was limited and should constrain the interpretation of the results.

Dynamic functional connectivity of the FPN may be a useful indicator of deficits in sustained attention in people with MS. Comparison between functional connectivity measurements at rest and during task-stimulated states may allow additional inferences which add value to standard task or resting-state fMRI individually. Further investigation of functional brain network dynamics in MS is warranted and may contribute to new mechanistic insights into cognitive dysfunction.

## Data Availability

Summary statistical data generated during and/or analysed during the current study are available from the corresponding author on reasonable request. Original datasets analysed are not publicly available to protect patient information.

## Supplemental

Supplemental information, Tables S1-5.

## Acknowledgements

We thank Prof Nadina Lincoln, Ms Cara Knight and Ms Holly Griffiths for help with recruitment; and Andrew Cooper for MRI scanning.

## Author Contributions

Conceived the study: RAD, DPA, CSC, TW. Performed image analysis: TW, DM. All authors discussed the results and contributed to the final manuscript.

