## Supplemental for "Fronto-parietal network dynamics to understand deficits in attention performance in multiple sclerosis"

**Supplemental Information**

***Sample Size Estimation***

We initially aimed to recruit 30 people with MS and 30 controls. This target sample size was arrived at by a power analysis based on an unpublished pilot study which included 8 people with MS and 8 controls. In order to demonstrate a significant group difference in measures with a large effect size at 80% power and with α=0.05, for a two-tailed independent samples t-test, a sample of 27 in each group was required. Acquisition of data from 3 additional participants per group was proposed to account for expected loss of data due to excessive motion or corruption (equivalent to an 11% expected rate of data loss), which gave us an estimated requirement of 30 participants per group.

The final sample sizes deviated from this target due to changes in the recruitment pathway mid-study. Since our actual group sizes were 37 vs. 23 for fMRI and 33 vs. 27 for structural tests, for the same parameters as above, our estimated power was 84% and 82%, respectively.

***MRI Acquisition Parameters***

- T1-weighted (FSPGR; FA=8°, matrix 256x256x156, voxel size 1x1x1mm, TE=3.17ms, TI=900ms, TR=8200ms, FOV=256mm, NEX=1).
- T2-weighted fluid attenuated inversion recovery (FLAIR; FA=111°, TE=120ms, TR=8000ms, TI=2250ms, matrix 512x512x46, voxel size 0.46x0.46x3mm, FOV=235.5mm).
- Diffusion MRI (dMRI; four b=0 volumes and 32 diffusion weighted volumes at b=1000, matrix 128x128x66, voxel size 2mm^3,^ TE=minimum, TR=8000ms).
- Functional MRI (fMRI; FA=80°, TE=36ms, TR=2200ms, matrix 64x64x37, voxel size 3.75mm^3^) at rest (9 minutes, 245 volumes) and during task performance (6 minutes, 165 volumes). During resting-state fMRI, subjects were instructed to close their eyes but stay awake and lie still.

***Brain Volumetrics***

The volume of white matter lesions was significantly greater in the MS group (7.77±8.87 mL) compared to controls (0.42±0.87 mL; t=3.95, p<0.001, Hedge’s g=1.04). Brain volumes were also significantly lower in the MS group (MS: 1014±104 mL, control: 1111±101 mL, t=3.47, p=0.001, Hedge’s g=0.92) and for both cortex (MS: 416±42 mL, control: 452±49 mL, t=2.92, p=0.005, Hedge’s g=0.77) and cerebral white matter separately (MS: 422±56 mL, control: 471±50 mL, t=3.35, p=0.001, Hedge’s g=0.88).

The total cortical volume of regions in the frontoparietal network (FPN) was significantly lower in the MS group (MS: 82.5±8.9 mL, control: 91.96±10.9 mL, t=3.58, p=0.001, Hedge’s g=0.95) but not when controlling for total brain volume (MS: 0.0815±0.0058, control: 0.0827±0.0049, t=0.78, p=0.44, Hedge’s g=0.21). The volume of the FPN was greater in the left hemisphere than the right (Figure 3c; paired-samples t-test across both cohorts; left: 423.6±4.7 mL, right: 409±4.4 mL, t=7.29, p<0.001, Hedge’s g=1.94).

**Supplemental Tables**

***Supplemental Table 1. Z-scores normalised to a reference sample and one-sample t-test.*** *For the ANT, no appropriate normative sample data were available. n/s = non-significant.*

|  | **Mean** | **SD** | **p** | **Source of Normative Data** |
| --- | --- | --- | --- | --- |
| **MSFC** | -0.72 | 0.66 | <0.001 | [12, 13] |
| **PASAT** | -0.72 | 1.14 | 0.001 | [12, 13] |
| **9HPT** | -0.58 | 1.13 | 0.003 | [12, 13] |
| **T25FW** | -0.06 | 0.55 | n/s | [12, 13] |
| **PSQI** | -0.05 | 0.74 | n/s | [14] |
| **MFIS** | 0.47 | 1.19 | 0.021 | [15] |
| **BDI** | -0.25 | 1.18 | n/s | [16] |
| **SDMT** | -0.67 | 1.06 | 0.001 | [17] |

***Supplemental Table 2. Paired-sample t-tests comparing static and dynamic connectivity measures during task performance to those at rest. FPN=frontoparietal network***

|  | t | p |
| --- | --- | --- |
| Multiple Sclerosis | | |
| FPN, Static Functional Connectivity | 0.731 | 0.469 |
| FPN, Dynamic Functional Connectivity | -2.627 | 0.012 |
| Whole-Brain, Static Functional Connectivity | 0.686 | 0.496 |
| Whole-Brain, Dynamic Functional Connectivity | -1.508 | 0.139 |
| Healthy Controls | | |
| FPN, Static Functional Connectivity | 1.807 | 0.075 |
| FPN, Dynamic Functional Connectivity | -6.381 | <0.001 |
| Whole-Brain, Static Functional Connectivity | 0.707 | 0.482 |
| Whole-Brain, Dynamic Functional Connectivity | -1.212 | 0.229 |

***Supplemental Table 3. Linear regression of task induced connectivity changes in the*** FPN ***against the ANT Alerting score across both groups, controlling for age, gender, education and head motion.***

|  | F(5,55) | p | R^2^ |
| --- | --- | --- | --- |
| Task-Induced Static Functional Connectivity Change | 9.150 | <0.001 | 0.234 |
| Task-Induced Dynamic Functional Connectivity Change | 1.087 | 0.302 | 0.019 |

***Supplemental Table 4. Linear regression of secondary attention performance measures against brain connectivity measures in the* FPN *in the MS group, controlling for age, gender, education and head motion.***

|  | F | p | R^2^ |
| --- | --- | --- | --- |
| Static Functional Connectivity | | | |
| SDMT | 0.031 | 0.860 | 0.001 |
| PASAT | 0.315 | 0.577 | 0.006 |
| Dynamic Functional Connectivity | | | |
| SDMT | 4.741 | 0.032 | 0.086 |
| PASAT | 4.667 | 0.035 | 0.082 |

SDMT, Symbol-Digit Modalities Test; PASAT, Paced Auditory Serial Addition Test.

***Supplemental Table 5. Comparison between MS and control groups on functional brain connectivity measures during task performance. FPN=frontoparietal network.***

| *Imaging Measure* | *F* | *p* | *R^2^* |
| --- | --- | --- | --- |
| *FPN* | | | |
| *Task Static FC* | *7.645* | *0.008 *** | *0.101* |
| *Task Dynamic FC* | *1.245* | *0.355* | *0.017* |
| *Whole Brain* | | | |
| *Task Static FC* | *0.004* | *0.951* | *0.004* |
| *Task Dynamic FC* | *0.416* | *0.521* | *0.010* |

* p<0.05

** p<0.01
